# Optimal use of COVID19 Ag-RDT screening at border crossings to prevent community transmission: a modeling analysis

**DOI:** 10.1101/2021.04.26.21256154

**Authors:** Joshua M Chevalier, Karla Therese L Sy, Sarah J Girdwood, Shaukat Khan, Heidi Albert, Amy Toporowski, Emma Hannay, Sergio Carmona, Brooke E Nichols

## Abstract

**Background:** Countries around the world have implemented restrictions on mobility, especially cross-border travel to reduce or prevent SARS-CoV-2 community transmission. Rapid antigen testing (Ag-RDT), with on-site administration and rapid turnaround time may provide a valuable screening measure to ease cross-border travel while minimizing risk of local transmission. To maximize impact, we developed an optimal Ag-RDT screening algorithm for cross-border entry.

**Methods:** Using a previously developed mathematical model, we determined the daily number of imported COVID-19 cases that would generate no more than a relative 1% increase in cases over one month for different effective reproductive numbers (Rt) of the recipient country. We then developed an algorithm- for differing levels of Rt, arrivals per day, mode of travel, and SARS-CoV-2 prevalence amongst travelers-to determine the minimum proportion of people that would need Ag-RDT testing at border crossings to ensure no greater than the relative 1% community spread increase.

**Findings:** When daily international arrivals and/or COVID-19 prevalence amongst arrivals increases, the proportion of arrivals required to test using Ag-RDT increases. At very high numbers of international arrivals/COVID-19 prevalence, Ag-RDT testing is not sufficient to prevent increased community spread, especially for lower levels of Rt. In these cases, Ag-RDT screening would need to be supplemented with other measures to prevent an increase in community transmission.

**Interpretation:** An efficient Ag-RDT algorithm for SARS-CoV-2 testing depends strongly on Rt, volume of travel, proportion of land and air arrivals, test sensitivity, and COVID-19 prevalence among travelers.

**Funding:** USAID, Government of the Netherlands

## INTRODUCTION

Severe acute respiratory syndrome coronavirus-2 (SARS-CoV-2), the causative agent of COVID-19, first emerged in Wuhan, China in late 2019. Aided by globalization, the novel coronavirus quickly spread to countries around the world. On the 11th of March 2020, COVID-19 was officially declared a global pandemic by the World Health Organization (WHO). In response to the outbreak, many countries implemented control measures on mobility, such as travel restrictions to and from affected countries, border closures, or quarantine-on-arrival.^1^

Once sustained community transmission is established within a country, there is little evidence that travel restrictions will continue to slow the epidemic as incidence is primarily driven by community spread.^3^ Some countries have had success containing community transmission and reducing the effective reproductive number (Rt) of SARS-CoV-2, producing a strong global impetus to prevent the importation of new infectious cases.^4–7^ This is particularly important as new, more infectious variants of SARS-CoV-2 begin to spread around the globe and risk circumventing progress on the distribution of effective COVID-19 vaccines.^8,9^ Effective travel related control measures are still needed to prevent the spread of old and new SARS-CoV-2 variants.

A key strategy to combat the COVID-19 pandemic is widespread accessible diagnostic testing.^10–12^ Today, countries around the world are implementing travel restrictions that require all international travelers to provide proof of a negative COVID-19 reverse transcription polymerase chain reaction (RT-PCR) test within 72-hours of arrival. While this is possible to implement in many high-income countries and for air travel, this is frequently not possible at land-border crossings, particularly in low- and middle-income countries (LMIC), due to frequent cross-border travel and resource constraints. Global partnerships have sought to boost testing capacity in Africa specifically by providing high-quality rapid antigen diagnostic tests (Ag-RDTs) that have been approved for emergency use by the WHO. In September 2020, the Access to COVID-19 Tools (ACT) Accelerator announced that 120 million Ag-RDTs would be made available for LMICs.^13^ Ag-RDTs are less costly than RT-PCR tests, do not require laboratory-based infrastructure, can be performed on-site by appropriately trained non-laboratory staff, and provide results within minutes, enabling decentralization of diagnostic testing.^12^

We aimed to develop a generalizable parsimonious algorithm for the use of Ag-RDTs at border crossings to determine the proportion of travelers that would need to be tested with Ag-RDTs to ensure international travel does not increase the baseline community transmission within a 1% margin of error. While such a strategy may provide greater benefit in settings with high volumes of land border crossings or limited RT-PCR testing capacity, the results of this analysis have broad implications for the use of Ag-RDTs in COVID-19 surveillance world-wide.

## METHODS

We developed a generalizable Ag-RDT screening strategy for use at border crossings with the goal of preventing a change in community transmission over a one-month period within the recipient country within a 1% margin of error compared to a baseline of zero imports. This algorithm was developed for differing effective reproductive numbers at time *t* (Rt) under varying levels of Ag-RDT sensitivity, number of daily travelers, and COVID-19 prevalence among travelers. Our algorithm determines the proportion of border crossings that would need to be tested per day with Ag-RDTs to prevent the relative 1% increase in transmission. We applied this algorithm to three case examples using real-world data on border crossings.

### Maximum allowable daily COVID-19 importations

To determine the number of daily infectious COVID-19 case imports that would lead to no more than a relative 1% change in community transmission over a one-month period compared to a baseline of zero imports, we used a COVID-19 SEIR model developed at the Biozentrum, University of Basel.^14^ This model has been used previously to make inferences about the spread of COVID-19 in various countries.^15^ To evaluate whether there was a linear relationship between the number of imported cases per day and community transmission, we exported the relative change in cases over a one-month period for increasing levels of daily SARS-CoV-2 importation (0, 10, 100,1000 infected persons per day) for four different levels of Rt (Rt=0.73, 0.93, 1.09, 1.45) for eight different countries (three of which were selected for further analysis). Model pre-set parameters for each selected country were used (15 November 2020) apart from the number of daily imports and Rt. Using these model exports, we created a linear equation (with R^2^=1) for each level of Rt that calculates the daily number of COVID-19 importations that would lead to a relative 1% increase in community transmission over the one-month period. The process was then repeated for all four levels of Rt.

### Algorithm Parameters

#### Number of International Arrivals

The number of international arrivals is expressed as the total number of people entering a country (by land or air) per day per 100,000 population of the host country. The number of international arrivals per 100,000 population of the host country varies substantially depending on the country, stage of the pandemic and levels of international travel.

#### Prevalence of COVID-19

True prevalence of people infectious with SARS-CoV-2 at a given point in time within a country is difficult to measure given the high incidence of asymptomatic infection and limited diagnostic testing capacity in many settings. Therefore, we simplified this parameter in our algorithm by varying COVID-19 prevalence (0.2%, 0.5%, 1%, 2%) among imports to model how a change in incidence would influence the Ag-RDT screening strategy.

#### Ag-RDT Sensitivity

Standard Ag-RDT sensitivity was assumed to be 80% in the base case, as determined through independent evaluations conducted by Foundation for Innovative New Diagnostics (FIND) on two WHO approved Ag-RDT tests being made available to LMICs.^16,17^ Due to variation in test performance results as well as to test the relative importance of test sensitivity (especially when testing asymptomatic individuals), we vary test sensitivity (50%, 80%, 90%) in our algorithm.

#### RT-PCR Testing

Airline travelers are required to present proof of a negative COVID-19 RT-PCR test prior to travel in many circumstances. A certain percentage of infections will be missed as a result of testing error or insufficient viral load. We have assumed that 12% of those infected with COVID-19 will not be detected through RT-PCR testing prior to air travel, based on a model simulation that found an 88% reduction in actively infectious travelers when tested with RT-PCR within 72-hours of travel.^18^ We have assumed that 100% of passengers traveling by air will have a negative RT-PCR test within 72-hours prior to travel and that 0% of passengers traveling by land will have a negative RT-PCR test within 72-hours prior to travel in this analysis.

### Interpretation and case examples

The model compared the number of undetected COVID-19 cases that would enter the country under the given conditions with the tolerable number of COVID-19 imports calculated for that level of Rt and determines the minimum proportion of arrivals that would need to be screened.

To illustrate how to interpret the algorithm results, we present results that apply the algorithm to South Africa, Germany and Australia, representing three different contexts. Arrivals to South Africa and Germany are composed of both land and air arrivals, with a majority of land in both cases. Australia represents an extreme scenario, having no shared land borders and nearly full control over community spread, providing an example of how an Ag-RDT screening strategy would work in an elimination context.^19^ Through these three case examples, we evaluate the effect of testing all land entries, all air entries, or a combination of both, assuming all air travelers are entering with proof of a negative COVID-19 RT-PCR test.

Data on international arrivals was obtained for the case examples from 2019 and 2020 for each individual country.^20–25^

## RESULTS

The minimum proportion of international arrivals required to be screened using Ag-RDTs at the border to prevent a relative 1% increase in community transmission over a one-month period depends on the total number of daily border crossings per 100,000 population of the recipient country, the prevalence of COVID-19 among arrivals, the proportion traveling into the country by land or air, and Ag-RDT sensitivity. As the Rt of the recipient country decreases, the proportion of international arrivals required to be tested increases. Border Ag-RDT screening alone is insufficient at high volumes of travel, when COVID-19 prevalence among international arrivals is high, and when Ag-RDT sensitivity is low (**Figure 1**). When the Rt is greater than one, Ag-RDT screening of border crossings is unnecessary at lower levels of travel volume and/or lower levels of COVID-19 prevalence among international arrivals. Ag-RDTs with higher sensitivities allow for a wider range of parameters in which a simple Ag-RDT screening strategy would be effective at preventing >1% increase in community transmission over a one-month period in the recipient country.

**Figure 1.**
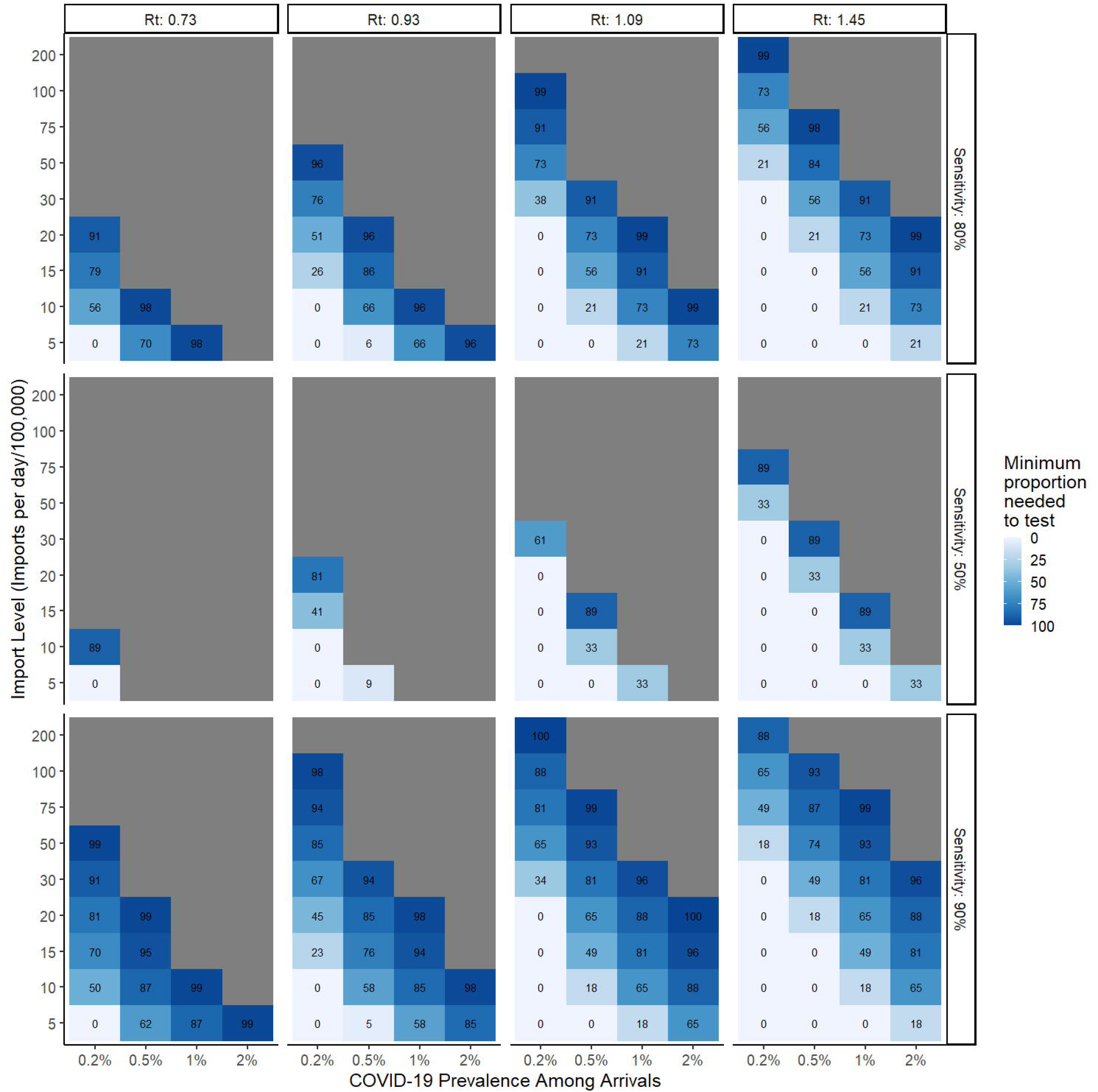
Impact of Ag-RDT test sensitivity on the minimum proportion of all arrivals (land and air) to be screened per day at the border using SARS-CoV-2 Ag-RDT tests. Varied by the reproductive number at time *t*, number of arrivals per day, and prevalence of COVID-19 among arrivals. The number in each cell represents the minimum proportion to be tested, while grey cells indicate Ag-RDT screening under the given conditions (even at 100%) is insufficient to prevent a relative 1% increase in community transmission in the host country over the course of one month.

Importantly, efficiency of the algorithm can be improved by focusing the use of Ag-RDT testing on the borders where arrivals do not have a negative RT-PCR test on arrival (e.g., through many land borders). When the vast majority of international arrivals into a country are, however, via air, then airports become the only logical place to implement border testing using Ag-RDTs. **Figure 2** illustrates the proportion of land border crossings that need to be screened with Ag-RDT when screening is only implemented at land crossings, **Figure 3** illustrates the proportion of air travelers that need to be screened with Ag-RDT when only implemented for air arrivals, and **Figure 4** illustrates the proportion of air travelers that would need to be screened in addition to full saturation of land border testing.

**Figure 2.**
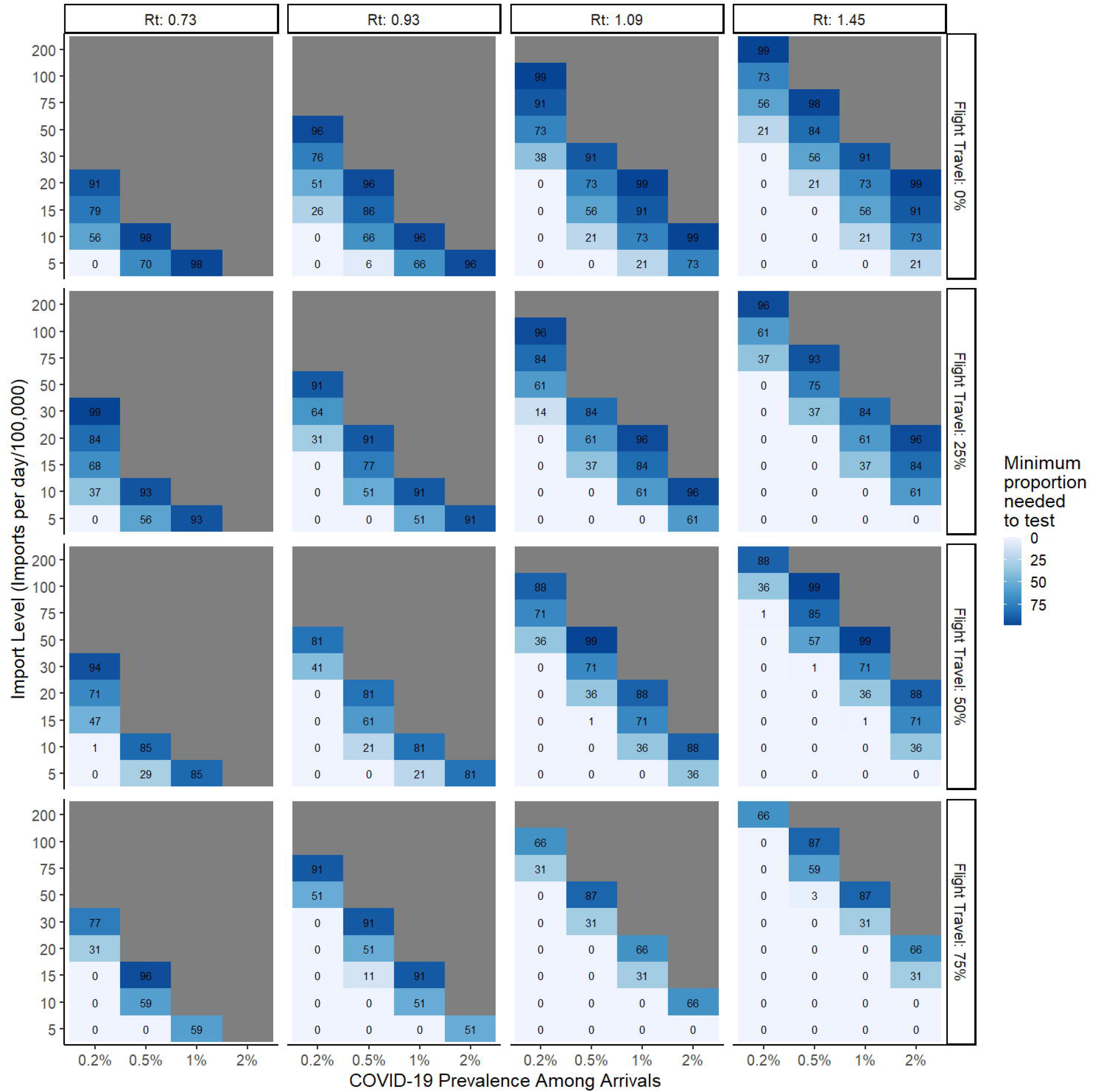
Impact of increasing proportion of flight travel on the minimum proportion of arrivals to be screened per day at land border crossings using SARS-CoV-2 Ag-RDTs when varied by the reproductive number at time *t*, number of arrivals per day, and prevalence of COVID-19 among arrivals. Ag-RDT sensitivity is set to 80%.

**Figure 3.**
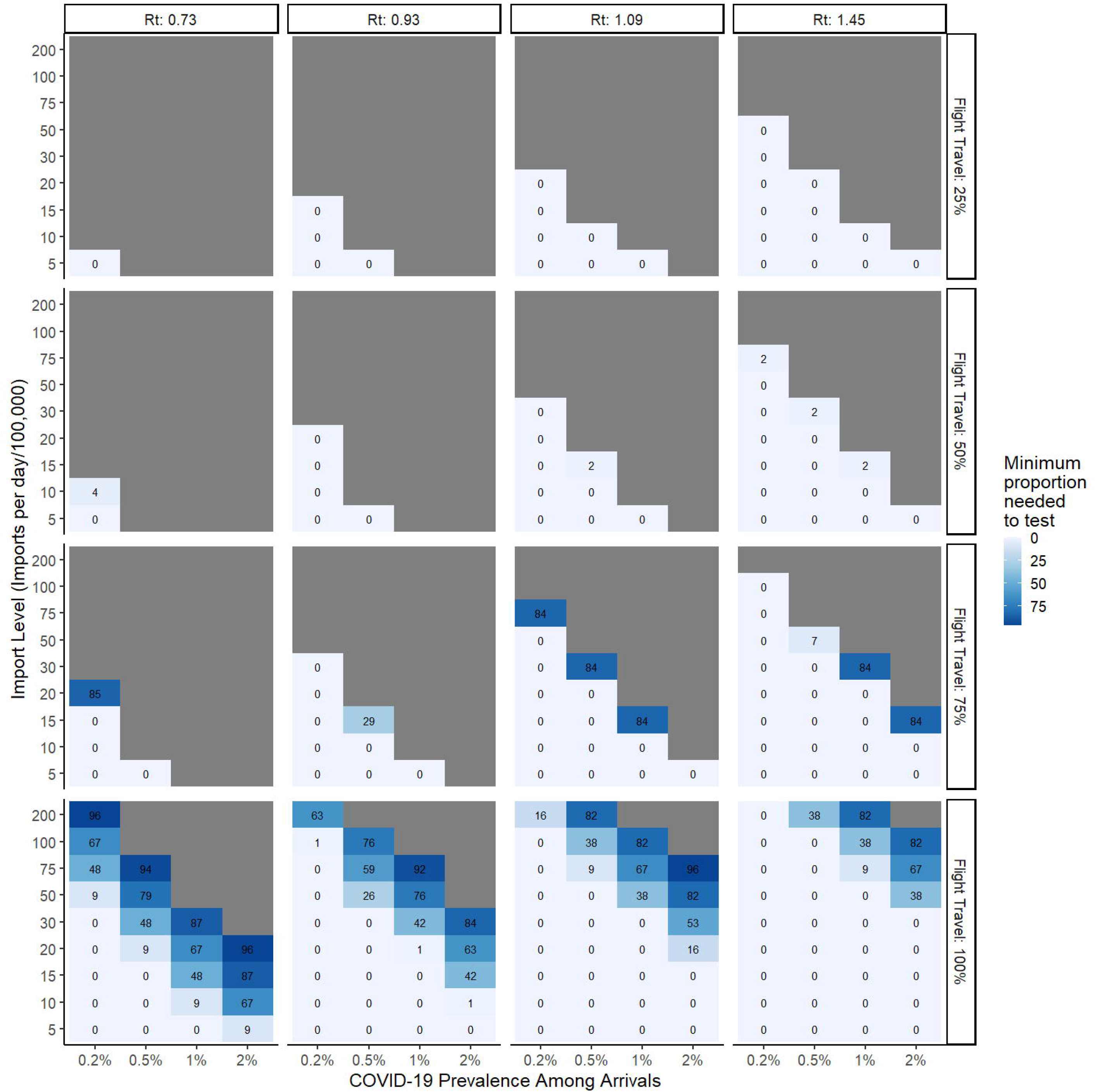
Impact of increasing proportion of flight travel when Ag-RDT screening is only implemented for flight arrivals on the minimum proportion of travelers to be screened with SARS-CoV-2 Ag-RDTs when varied by reproductive number at time t, number of imports per day and SARS-CoV-2 prevalence. Ag-RDT sensitivity is set to 80%.

**Figure 4.**
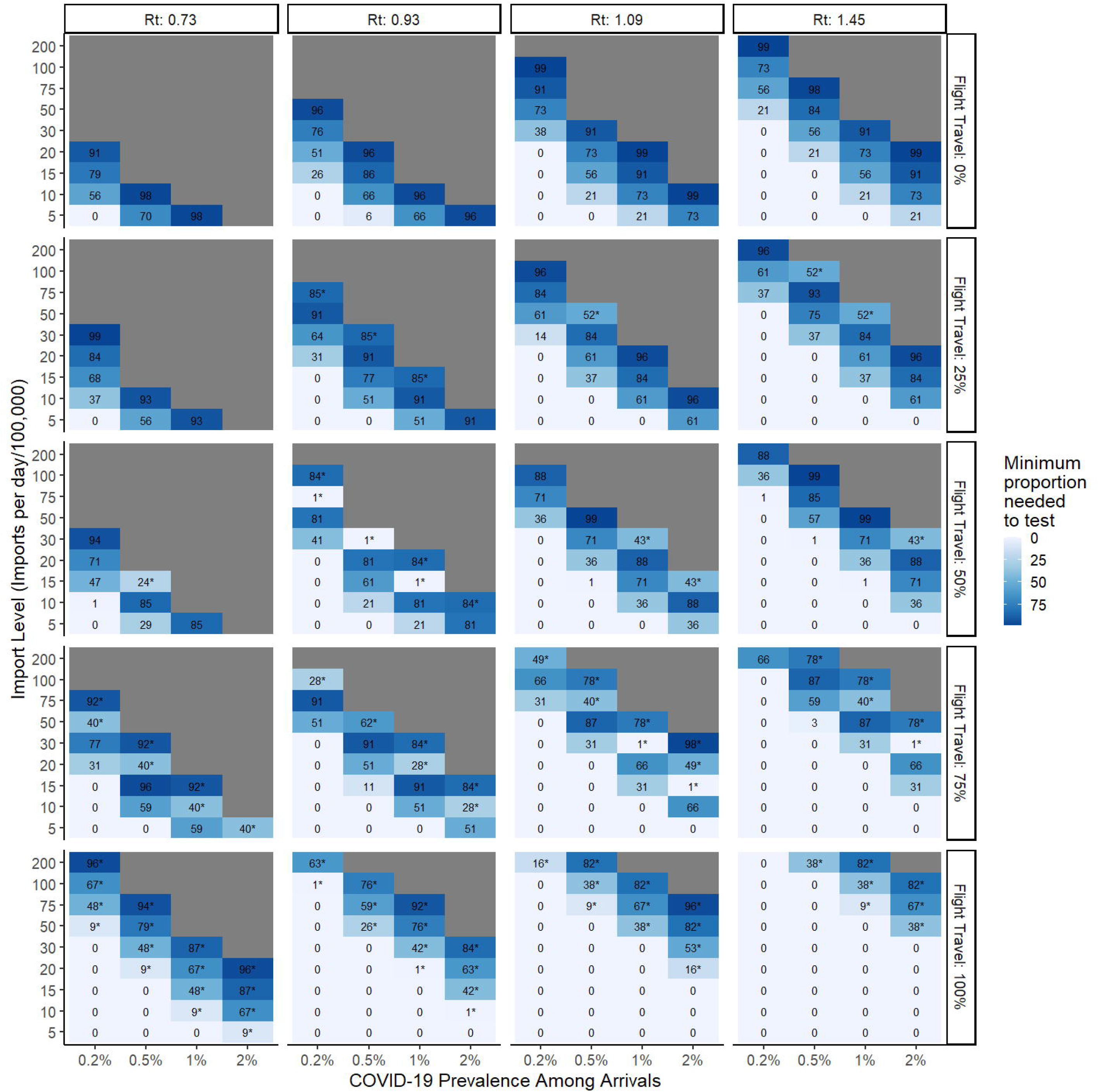
Impact of increasing proportion of flight travel when Ag-RDT screening is first fully saturated at land borders and additionally implemented for flight arrivals. Cells with an asterisk (*) represent the additional proportion of flight arrivals to be screened with SARS-CoV-2 Ag-RDTs following full saturation of land testing. Cells without an asterisk correspond to the minimum proportion of land arrivals required to be tested (Figure 2).

### Case examples

#### South Africa

International travel into South Africa was reduced by nearly 90% for most of 2020 as compared with 2019 but, increased during the fourth quarter. In the fourth quarter of 2020, composed of both South African residents and non-resident tourists, the majority of arrivals entered by land (72%), while 28% entered by air.^20–22^ The average number of international arrivals per day including both land and air was 16 per 100,000 population during the fourth quarter of 2020. Under the given travel conditions, in the base case with a test sensitivity of 80%, Ag-RDT screening of land arrivals would be sufficient for low COVID-19 prevalence when the Rt is low (<1) and for high COVID-19 prevalence when the Rt is high (>1) (**Figure 2**). Screening of land and air arrivals increases the sufficient range only slightly under the given conditions (**Figure 4**).

#### Germany

Tourist travel to Germany was down nearly 85% during the year 2020 as compared with 2019. Restricted to non-resident foreign tourist arrivals, the average number of daily arrivals in quarters three and four of 2020 was 40 per 100,000 population.^23^ We applied the breakdown for mode of entrance of tourist arrivals from 2019, which consisted of 59% arrivals by land and 41% by air.^24^ Under these conditions, in the base case with a test sensitivity of 80%, Ag-RDT screening of land arrivals is only sufficient at low levels of COVID-19 prevalence and/or at moderate to high levels of Rt (**Figure 2**). Screening of land and air arrivals only increases the effectiveness of the algorithm marginally compared with land arrivals alone (**Figure 4**).

#### Australia

Australia has experienced the most drastic reduction in incoming travelers throughout 2020, seeing a decrease of 99% compared with 2019.^25^ All international arrivals are assumed to enter Australia exclusively by air with a negative COVID-19 RT-PCR test within 72-hours of travel and are composed of both residents and non-residents. The average number of daily arrivals in quarters three and four of 2020 was 3 per 100,000 population, while the tolerable threshold of infectious imports per day was less than one imported infection for every level of Rt. Based on these conditions the Ag-RDT screening strategy is only sufficient for low level COVID-19 prevalence when the Rt is >1 and insufficient in all other scenarios to prevent the relative 1% increase in community transmission. Generally, in situations where there is no longer community transmission, Ag-RDT screening at borders must be combined with quarantine measures to prevent re-seeding of the epidemic.

## DISCUSSION

The effectiveness of a SARS-CoV-2 Ag-RDT screening strategy at border crossings depends largely on the current state of the COVID-19 pandemic within the recipient country (represented by Rt), the number of daily international arrivals into the recipient country, the proportion entering by land versus air (given the negative RT-PCR required within 72-hours of air travel), the expected COVID-19 prevalence among arrivals, and the sensitivity of Ag-RDTs. As the Rt in the recipient country decreases, the proportion of border crossings that need to be tested to prevent the relative 1% increase in community transmission increases. As travel volume and COVID-19 prevalence increase, so does the proportion required to test. At high travel volume and COVID-19 prevalence, the Ag-RDT screening strategy becomes ineffective on its own, especially at lower levels of Rt. In these situations, 100% of arrivals would need to be tested, and then further supplemented by quarantining and additional testing. Border Ag-RDT testing would no longer be required to prevent a relative 1% increase in community transmission when Rt is high and when travel volume and COVID-19 prevalence are low. In such situations, Ag-RDTs could be better reallocated for other uses and utilized to contain internal community spread.

We demonstrate several scenarios in which the proportion of international arrivals that would require an Ag-RDT test ranges from 0% to 100%. This could be implemented in a number of ways to increase impact or possibly reduce the total proportion of people needed to test. In particular, these strategies could include 1) focusing Ag-RDT testing at land crossings specifically or 2) testing 100% at some borders where prevalence is higher, and 0% others; 3) a combination of 1 and 2. A COVID-19 Ag-RDT border screening strategy is largely context dependent and may need to be implemented alongside additional non-pharmaceutical interventions (NPIs) to maximize utility. Our case examples of South Africa and Germany have a range of scenarios where an Ag-RDT screening strategy would be effective on its own. The effectiveness could be further enhanced with a combination of additional NPIs in the scenarios where 100% of border screening using Ag-RDTs is insufficient to prevent a relative 1% increase in community transmission over the course of one month. Additionally, focusing testing first at land borders, followed by air arrivals would increase the range of scenarios where testing alone is sufficient.

Australia on the other hand, represents an extreme scenario in which community spread of COVID-19 was eliminated at the time of this study and therefore remains highly sensitive to the importation of new cases. In Australia Ag-RDT screening of travelers on arrival is not sufficient in the majority of circumstances, even when those travelers have a prior negative RT-PCR test. The tolerable threshold of infectious imports in all scenarios was <1 case of SARS-CoV-2 per day, meaning no amount of single point testing is theoretically capable of catching all infectious imports. It thus becomes necessary to accompany testing with quarantine-on-arrival to prevent substantial impact on community transmission.

To the best of our knowledge, this is the first study to examine the role of Ag-RDTs at border crossings, incorporating both land and air imports. A limited number of studies have evaluated the effect of RT-PCR testing on COVID-19 importations.^27,28^ *Russel et al*. modeled the effect of travel restrictions on the importation of COVID-19 cases and their contribution to local incidence, finding that travel restrictions have little impact on local epidemics under conditions of high community transmission, but may play an important role when COVID-19 incidence is low within the country.^3^ Countries like New Zealand and Australia would not have been able to eliminate community spread without strict border control and containment measures.^29,30^ Modeling of New Zealand’s current arrival protocols with variations concluded that 14-day quarantine on arrival with two consecutive negative RT-PCR tests is the most effective regimen to maintain elimination in the country.^30^ These studies suggest testing alone is unlikely to completely eliminate the risk of international arrivals seeding new outbreaks. The role of Ag-RDTs in such strategies ultimately depends on the goal of each country. Our analysis is the first attempt at quantifying the amount of testing necessary for a point Ag-RDT screening strategy at border crossings, evaluated against a predetermined tolerable increase in community transmission. Our generalizable algorithm can be used directly by high-, low- and middle-income countries to help inform border testing policy and resource allocation.

Our analysis comes with important assumptions and limitations. First, due to the nature of COVID-19 prevalence, serology studies, and differing testing algorithms by country, determining the true incidence of COVID-19 at any given instance is not feasible. To address this, we have assumed a maximum incidence of 2%, and provide an increasing range up to this point. Second, our model output directly relies on the 1% threshold calculated for each Rt in each individual country. However, the 1% margin of error threshold could theoretically be modified to re-define what a tolerable increase in community transmission from outside imports would be for a given country. Third, our algorithm evaluates the benefit of a single point/once-off Ag-RDT screening strategy on its own and was not analyzed in combination with follow-up testing or other mitigation measures that could enhance its effect, as this falls outside the scope of our analysis. The decentralized nature of Ag-RDT testing, while beneficial, may prove difficult to implement alongside additional strategies, such as contact tracing—particularly in limited resource settings. Finally, if the goal of a given country is to prevent any SARS-CoV-2 importation, Ag-RDT screening is likely to be insufficient and would need to be implemented in combination with quarantine-on-arrival measures.

Optimal allocation of Ag-RDTs to border crossings depends strongly on Rt, volume of travel, proportion of land and air arrivals, test sensitivity, and COVID-19 prevalence among travelers. When travel and incidence are low, implementing a border screening strategy could be beneficial, but when both are high a country may consider reallocating their Ag-RDTs to other testing programs within their country. As with many decisions in this pandemic, the optimal border testing algorithm for a given country will depend on the goal of the testing strategy.

## Data Availability

All data utilized in this manuscript has been referenced and is publicly available.

